# Development and Validation of a Bedside Scale for Assessing Upper Limb Function Following Stroke: A Methodological Study. [version 1; peer review: awaiting peer review]

**DOI:** 10.1101/2024.01.29.24301976

**Authors:** Dhaval Pawani, Abraham M. Joshua, Akshatha Nayak, Vijayakumar Palaniswamy, Prasanna Mithra, Ashish John Prabhakar, Sampath Kumar Amaravadi

## Abstract

**Background:** Numerous tools are available for evaluation of upper limb (UL) functions among stroke survivors. Despite the excellent psychometric properties, many require considerable amount of time, are resource-intensive, and often impractical for bedside evaluation.

**Objectives:** To develop and concurrently validate a simple, resource-efficient, and time-efficient bedside tool for evaluating UL function in stroke survivors.

**Methods:** Relevant literature review was carried out to conceptualize and define the theoretical framework of day-to-day UL movement tasks. Subsequently, an item pool of 18 UL movements was developed. A mini-Delphi method was employed to verify content validity. During the iterative rounds, 18-items were revised and refined to a 12-items scale. The final bedside upper limb evaluation tool (BUFET) scale underwent concurrent validation by correlating the scores with Wolf Motor Function Test (WMFT) scores using Spearman’s correlation coefficient. Internal consistency was evaluated through Cronbach’s alpha.

**Results:** Concurrent validity and internal consistency of the scale were supported by a high correlation coefficient (r = 0.937; p<0.001) with WMFT and high Cronbach’s alpha (0.948).

**Conclusions:** Newly developed BUFET was found to be a valid and reliable bedside tool in the evaluation of upper limb functions and can be administered in a resource and time-efficient manner.

## Introduction

The daily activities of individuals with stroke are significantly influenced by the upper limb (UL) and hand function.^1^ Evidence from several studies suggested that 85% of stroke survivors suffer UL and hand impairments.^2–5^ In particular stroke survivors with middle cerebral artery infarction have been associated with muscle weakness,^6^ inability to control all UL segments in space and time (inter-joint coordination),^7,8^ difficulty in grasping and holding an object, and reduced ability to independently move individual fingers.^3^ There is significant evidence to suggest that these UL impairments contribute to loss of UL function, loss of independence in activities of daily living, and impaired quality of life.^9,10^ The presence of these diverse motor impairments a few weeks after a stroke can predict future UL function.^3^ Therefore, evaluation of UL function is critical in day-to-day stroke rehabilitation.^11^

Evaluation of UL functional movements following stroke has been performed by several types of tools ranging from observer-based scales, instrumented tests, and self-reported questionnaires.^12^ Some of the commonly reported reliable and valid performance assessment tools to quantify UL function in stroke survivors include the Fugl-Meyer Assessment of Upper Limb (FMA-UL), Wolf Motor Function Test (WMFT), Action Research Arm Test (ARAT), Box and Block Test (BBT), Nine Hole Peg Test (NHPT).^12–14^ Although FMA-UL has been reported to have the highest level of psychometric and clinometric properties, it does not evaluate functional arm and hand movements.^15^ FMA-UL mainly evaluates body function and structures as per the international classification of functioning (ICF) framework. In addition, FMA-UL and ARAT are noted to exhibit some overlap in their assessment of UL function, suggesting that they may not be entirely distinct in their evaluations of UL capabilities.^16^ Clinical tools such as ARAT, WMFT, BBT, and NHPT predominantly measure grasping and displacement movements of different object sizes with less emphasis on gross movements.^17,18^ ARAT involves a subjective scoring method, with poor definitions of the test item positioning and time allocation for each item.^14,19^ Currently, there is no agreement on the selection of any particular tool for a particular individual with a stroke.^3^

Despite the excellent psychometric properties of FMA-UL, WMFT, and ARAT, all these tools require a considerable amount of time and are resource-intensive due to their comprehensive nature and need for manual administration.^19^ In particular, FMA-UL requires longer than 30 minutes to complete the test and needs material resources and/or tools including a standardized chair and/or desk to execute the same.^20^ Furthermore, administration of these said measures can be exhaustive, cumbersome, and often impractical for bedside evaluation. The time taken to administer a tool significantly influences its probability of regular usage in clinical practice. Hence, tools that take a quicker time are more likely to be utilized.^3^

Emerging evidence suggests that the inclusion of non-contact gesture movements (e.g., salute and hand waving gesture) and contact-grasping (e.g., grasping a small glass) would strengthen the representativeness and comprehensiveness of evaluation of UL movements of daily life.^2,21^ In addition, analysis of gesture and grasp movements can demonstrate task-specific and impairment-specific characteristics.^21^ In line with that, we propose a conceptual framework for the clinical utility of day-to-day movement tasks such as hand gesture movements, grasping movements,^22^ and rhythmic finger tapping^23^ in evaluating the UL function in stroke. Although previous studies have quantified the impairments in hand gestures, grasping, and finger tapping, these studies primarily employed expensive quantifiable technology to investigate such as wearable gloves for hand gesture recognition, and ultrasound-based motion analyzer for kinetic and kinematic analysis of grasping.^22,24,25^ Evidence suggests that the finger-tapping test is a useful tool in predicting recovery in stroke survivors.^26,27^ Currently, there is no simple, qualitative, resource, and time-efficient tool that could be quickly administered at the bedside to evaluate UL function in stroke survivors.

Therefore, the primary aim of the present study was to develop a bedside tool based on day-to-day movement tasks that can be administered with ease, accuracy, minimal time consumption, and less exhaustion to measure the UL function following stroke. The secondary aim was to assess the concurrent validity of the new bedside tool.

## Methods

The study comprised of 2 phases 1) scale development and content validity verification 2) concurrent validity determination with WMFT. After receiving approval from the Research and Institutional Ethics Committee of Kasturba Medical College, Mangalore (IEC KMC MLR 1/2022/15) on 19/01/2022, a dual-phasic study containing qualitative and cross-sectional elements was undertaken in teaching hospitals affiliated with Kasturba Medical College, Mangalore, from February 2022 to January 2023. The study adhered to the ethical principles of the Declaration of Helsinki for research involving human participants. The qualitative phase included tool development, whereas the cross-sectional phase focused on tool validation. Purposive sampling was implemented for participant recruitment during cross-sectional phase. The study included adult participants (>18 years of age) diagnosed with primary infarction/hemorrhagic stroke and hemiparesis of the upper limb (UL). Exclusion criteria of the study were i) other neurological disorders, ii) severe cognitive deficits (Montreal Cognitive Assessment score < 24), iii) perceptual dysfunctions, and iv) pre-existing UL musculoskeletal conditions affecting testing.

### Scale development

The theoretical conceptualization and development of the new Bedside Upper Limb Evaluation Tool (BUFET)© was guided by AMJ. Initially, the research team identified 18 simple day-to-day movement tasks (Table 1 and Table 2) as potential scale items through a comprehensive review of relevant literature. The initial 18-items scale comprised of tasks such as UL and hand gesture movements, grasping movements, and finger tapping. All the identified movements require coordinated function of the shoulder, elbow, wrist, hand, and fingers.

**Table 1.** Items excluded with Delphi method.

| Item | Expert's rationale for exclusion |
| --- | --- |
| 15. Elbow flexion- Break test for strength examination | Excluded as this item focuses solely on evaluating the muscle strength rather than priority hand function. Also, its inclusion would necessitate further evaluation of multiple muscles to validate strength examination. |
| 16. Make a ring by opposing thumb & index- Examiner breaks to check the strength | Excluded due to duplicity, given its similarity to the action of "Gesture 3". |
| 17. Functional position of hand | Excluded as the scoring can be difficult and may resemble the typical hand posture observed in many stroke individuals. |
| 18. Elbow extension- Break test for strength examination | Excluded as this item focuses solely on evaluating the muscle strength rather than priority hand function. Also, its inclusion would necessitate further evaluation of multiple muscles to validate strength examination |

**Table 2.** Content validation for bedside upper limb functional evaluation tool.

| Items | E-1 | E-2 | E-3 | E-4 | E-5 | E-6 | Remarks | Included/ Excluded |
| --- | --- | --- | --- | --- | --- | --- | --- | --- |
| 1. Salute | 5 | 5 | 5 | 5 | 5 | 5 |  | Included |
| 2. Hold the nose | 4 | 5 | 5 | 4 | 3 | 4 |  | Included |
| 3. Hand Waving | 5 | 5 | 5 | 5 | 4 | 5 |  | Included |
| 4. Make a claw/ hook | 1 | 1 | 1 | 1 | 1 | 1 | Claw being a deformity is misfit as a component | Excluded |
| 5. Grip the examiner's fingers | 5 | 5 | 5 | 5 | 5 | 4 |  | Included |
| 6. Oppose and maintain the contact of thumb and little finger | 5 | 5 | 5 | 5 | 5 | 4 |  | Included |
| 7. Point the index finger upwards with wrist in extension | 4 | 5 | 5 | 5 | 5 | 5 |  | Included |
| 8. Clockwise and anticlockwise stirring action of wrist | 4 | 5 | 5 | 4 | 4 | 5 |  | Included |
| 9. Snapping action of the fingers | 4 | 5 | 5 | 4 | 4 | 4 |  | Included |
| 10. Hold the examiner's finger using the thumb and index finger | 5 | 5 | 5 | 4 | 4 | 3 |  | Included |

**Table 2.** *Continued*
| Items | E-1 | E-2 | E-3 | E-4 | E-5 | E-6 | Remarks | Included/<br>Excluded |
| --- | --- | --- | --- | --- | --- | --- | --- | --- |
| 11. Gesture a scissoring action using the index and middle finger | 5 | 5 | 5 | 5 | 5 | 5 |  | Included |
| 12. Interlacing of Fingers | 5 | 4 | 3 | 4 | 1 | 1 | A representation of abduction and adduction of fingers is covered in the “scissoring action” hence removed due to similarity | Excluded |
| 13. Gesture the number 3 using middle, ring, and little finger | 5 | 5 | 5 | 5 | 5 | 3 |  | Included |
| 14. Finger tapping | 4 | 5 | 5 | 5 | 5 | 5 |  | Included |

Note: E = Expert.

### Content validation

A mini-Delphi consensus method was implemented to achieve content validity, involving a series of rounds with expert consensus panel comprised of 6 clinical researchers with a minimum of 15 years of experience in specialized neurological clinical practice. The iterative method of mini-Delphi technique, as outlined by Hasson et al., involves multiple rounds of communication aimed at achieving consensus. The selection of experts ensured that relevant expertise and experience were present to provide valuable insights and feedback on the questionnaire items. In each round, the experts were provided with structured questionnaires, along with detailed instructions on how to provide feedback. The process allowed for anonymity, reducing the influence of dominant individuals and facilitating open expression of opinions.^28^ The iterative nature of the method allowed for structured communication among the experts, facilitating the exchange of opinions and feedback effectively.^29^ The expert panel included 3 neurologists, 2 physiotherapists, and 1 occupational therapist, ensuring a diverse range of perspectives in the evaluation process. This diversity contributed to consensus building among the panellists regarding the content and validity of the questionnaire items.

In the first round, experts individually reviewed the 18-item scale via email, maintaining anonymity for unbiased feedback. Subsequently, in the second round, consensus was reached to exclude 4 non-relevant items (item # 15 to 18, Table 1). The remaining 14-item scale (Table 2) underwent further iterative feedback rounds.

In the subsequent third round, experts provided critical feedback on the 14-item scale, leading to revisions based on consensus. This structured approach culminated in the development of a finalized 12-item BUFET, entering subsequent validation phases. The finalized 12-item BUFET, including rating scores, is available as an underlying data from https://doi.org/10.17605/OSF.IO/UFHK5.

### Concurrent validation

After developing the final version of scale, concurrent validation process was initiated with purposive sample of 25 stroke survivors. This phase involved recruiting 25 stroke survivors meeting inclusion criteria against the estimated minimum sample size of 20. The study participant characteristics are depicted in Table 3. All the participants were administered with BUFET and Wolf Motor Function Test (WMFT) randomly with a one-hour interval between the two tests. Evidence indicates that WMFT comprises 15 timed task-performance items that can assess functional ability with excellent reliability. The correlation between BUFET and WMFT scores of all 25 participants were determined using Spearman’s correlation coefficient method.

**Table 3.** Demographic and clinical characteristics of the participants.

| Characteristics | Total (N=25) |
| --- | --- |
| Gender- female/male | 8/17 |
| Mean age in years (SD) | 60.6 (9.55) |
| Lesion Location- Supratentorial (%) | 25 (100%) |
| Lesion Type-Infarction/hemorrhagic | 21 (84%)/4 (16%) |
| Side of Involvement-left/right | 9/16 |
| Mean MoCA/30 (SD) | 26.96 (1.65) |
| Mean WMFT/75 (SD) | 52.64 (12.75) |
| Mean BUFET/48 (SD) | 34.36 (7.97) |

Note: SD = standard deviation, % = percentage, N = number.

### Outcome variables

The WMFT includes 15 timed tasks with each item rated on a six-point functional ability scale, assessing effort, smoothness, and overall quality. At the outset, the scale tests the unaffected side, followed by the affected limb, and generally takes 30-35 minutes to complete the evaluation. Evidence suggests that WMFT exhibits excellent test-retest reliability (r=0.95) and strong inter-rater (ICC=0.93) and intra-rater (ICC=0.97) reliability. Required materials for WMFT include a standardized table, chair, box, 12-oz beverage can, 7” pencil with six flat sides, 2” paper clips, lock and key, face towel, and basket.^30–32^

### Statistical analysis

Statistical Package for Social Sciences (SPSS, Version 25.0, released 2017. IBM Armonk, NY: IBM Corp) was used for data analysis. The concurrent validity of BUFET was assessed by correlating the scores against those of WMFT using Spearman rank correlation analysis. The internal consistency of the tool was analyzed by obtaining Cronbach’s alpha.

## Results

### Content validation

The principal investigator (PI) organized an initial 18-item scale focusing on essential day-to-day movement tasks including gestures, grasping, and finger tapping (Table 1 and Table 2). This scale was subsequently revised to 14 items based on recommendations from the six subject experts who participated in the study (Table 2). Removal or modification of scale and/or its grades was considered if at least two subject experts rated a score of 2 or below on a 5-point Likert scale for that item. According to this specified criterion, two more items were eliminated, as shown in Table 2.

A closing agreement from the subject experts on the revised 12-item scale was carried out and the finalized BUFET comprised of 7 gesture items, 3 grasping or gripping items, 1 item for wrist movement, and 1 finger tapping.

### Participant characteristics

A total of 25 participants (17 males, 8 females), with a mean age of 60.6 years, participated in the concurrent validation study. All participants suffered supratentorial infarction (84%) or haemorrhagic type (16%) of stroke (Table 3).

### Correlation analysis

Correlation analysis was utilized to confirm the concurrent validity of the proposed BUFET scale by comparing them to WMFT which was used as reference standard. The normality of BUFET scores suggested a normal distribution and WMFT scores revealed absence of normal distribution. Since one of the variables was not normally distributed, Spearman rank method was used for correlation coefficient analysis. The results of analysis indicated a high significant correlation coefficient (r = 0.937; p < 0.001) as presented in the Figure 1. Additionally, the BUFET demonstrated high internal consistency, as reflected by a Cronbach’s alpha value of 0.948.

**Figure 1.**
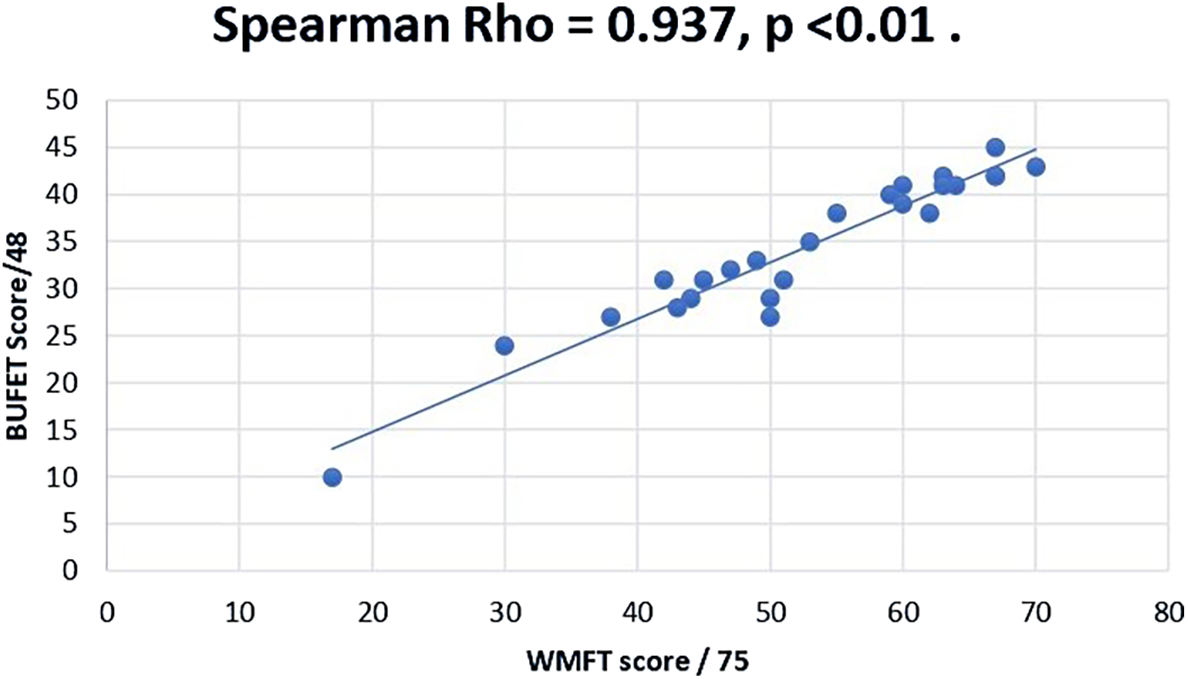
Correlation between BUFET and WMFT Scores. BUFET: Bedside Upper Limb Functional Evaluation Tool; WMFT: Wolf Motor Function Test.

## Discussion

In the current study, evidence is provided for the contention that day-to-day movement gestures, grasping activities, and rhythmic wrist and finger movements constitute a significant tool for the evaluation of UL function in stroke survivors. Previous research primarily focused on investigating the therapeutic efficacy of gesture, grasping, and finger-tapping movements to enhance UL function using quantitative and expensive methods.^2,22^ Nonetheless, there is an apparent significant gap in the literature regarding the development and validation of a qualitative bedside tool utilizing daily gesture, grasping, and finger tapping movements for evaluating UL function in stroke survivors. Existing tools in assessment of shoulder, elbow, wrist, and hand motor impairment require specific materials, training, and excessive amount of time. Consequently, a simple, inexpensive, resource (no resources) and time-efficient (<10 mins) Bedside Upper Limb Evaluation Tool (BUFET) was developed and validated with methodological study design. Such a qualitative bedside tool that can be implemented in clinical, research, or home settings could be a critical component in stroke rehabilitation.

Evaluation of UL and hand function is pivotal for the comprehensive rehabilitation of stroke survivors. The newly developed BUFET serves as a qualitative instrument that can be efficiently administered at the bedside. This tool facilitates the observation of intricate patterns in shoulder, elbow, hand, and wrist movements during the execution of gestures, grasping, and fine finger movements. According to Michael Roth, symbolic hand gestures are predominantly upper arm and hand movements, conceptualized as originating from ergotic hand movements associated with object manipulation and epistemic hand movements related to sensing activity.^33^ In general, hand orientation assumed by the grasping hand depends on the initial hand position, location, shape, and orientation of the object to be grasped.^34,35^ However, stroke survivors often exhibit impaired gesture imitation, influencing the performance of specific arm and hand segments (limb apraxia).^21^

Kinematic studies have indicated that complex hand gestures and grasp movements in stroke survivors are associated with altered joint rotation patterns, hand orientations, and impaired inter-joint coordination of grasp and twist.^2,7^ Evidence also suggests that impaired hand gesture imitation is linked to posterior lesions in the left inferior parietal lobule (LIPL) and temporal-parietal-occipital junction (TPOJ), while impaired finger gesture imitation is associated with lesions in the inferior frontal gyrus (IFG).^36,37^ These brain regions are responsible for motor planning, coordination, and the integration of sensory-motor information.^7^

Consequently, prompt qualitative movement analysis of gestures, grasping, and fine finger movements using BUFET at the bedside empowers the examiner to raise clinical suspicion regarding the potential location of brain lesions in stroke survivors. This tool facilitates early and timely interventions. In particular, the qualitative assessment focused on UL motor function can reveal distinctive patterns that correlate with the left inferior parietal lobule (LIPL) and temporal-parietal-occipital junction (TPOJ), enabling differentiation between stroke groups and offering valuable insights into the functional abilities of stroke survivors. Furthermore, it may provide indications of specific brain lesion types, distinguishing between posterior (LIPL) and anterior (IFG) lesions.

The BUFET covers a wide range of UL, hand, and finger movements. The first component-salute evaluates the ability to produce a movement pattern away from the typical attitude of the affected limb.^38^ The second and the third components (holding the nose and hand waving, respectively) help to assess the quality of control of shoulder flexors and external rotators which are reported to be considerably impaired among stroke subjects.^39^ Also, the second item (holding the nose) reflecting hand-to-mouth function is reported to be a significant method to evaluate UL in subjects with stroke.^40^ The functional ability of the intermediate joint (i.e., elbow), to achieve complete flexion is also tested in the first and second components while the elbow extension is tested in the third component. Levin et al emphasized that the movement amplitudes at the shoulder and elbow joints were significantly impaired during the excursion of the hemiparetic arm.^35^ In particular, during reach-out tasks, effective shoulder movements with inter-joint coordination are paramount.^41^ Hence, hand waving has been included as a third component to evaluate shoulder function.

Wrist circumduction movement is usually described as flexion-extension motion in function of radio-ulnar deviation. Most daily activities of life can be performed through an arc from 10° flexion to 35° extensions. Evidence suggests that static flexion posture of finger significantly influences wrist circumduction with a linear relationship between wrist and finger movements.^42^ This phenomenon is particularly implicated in wrist drop observed among people with unilateral stroke. A Rasch model analysis of the psychometric properties of wrist and hand subscales of FMA-UL in stroke reported a higher/large positive factor loading (0.846) for wrist circumduction representing the unidimensional construct of UL and hand motor function.^43^ Thus, inclusion of clockwise and anti-clockwise stirring action of the wrist as one of the items demonstrated that BUFET is unidimensional. Furthermore, reduced selectivity of the muscles that control isolated index finger extension, a deficit in the ability to perform isolated finger extension, has also been studied.^44^ Hence, inclusion of pointing the index finger upwards with the wrist in extension is considered significant.

Maximum grip force is generated with the wrist held in extension,^22^ and lack of recovery of grasp efficiency may suggest the inability of the descending pathways to control the distal muscles.^45^ Additionally, the radial aspect of the hand plays a significant role in fine motor tasks such as gripping, which require greater dexterity and strength. Liu et al. stated that the thumb, index, and middle fingers that are controlled by the radial aspect of the hand are more prone to impairment compared to other fingers.^46^ Due to these reasons, the evaluation of the grip strength through the radial aspect is considered.

The opposition of thumb is essential for daily activities like picking up small objects. The opposition was noted to be reduced in stroke subjects when compared to healthy.^47^ Nijland et al. reported that the ability to extend the finger within 72 hours post-stroke can predict functional recovery in the hemiplegic arm at 6 months.^48^ While attempting to move a specific digit, inappropriate contractions of muscles in other digits were noted among stroke subjects.^44^ Prior research also stated an increased level of motor impairment with ulnar fingers i.e., middle, ring, and little finger.^49^ Since specific digit(s) movements are likely to be impaired in stroke subjects pointing the index finger upwards and gesturing the number “3” are included.

Studies have reported impaired thumb movements and reduced velocity in finger flexion movements among stroke subjects.^50^ To evaluate thumb and middle finger control, the snapping action of the finger that requires quick flexion of the middle finger against the thumb is selected as a component. In addition, finger abduction/adduction was reported to be greatly impaired compared to flexion/extension.^44^ Thus, BUFET included the scissoring action of the index and middle fingers as a test item to evaluate the finger abduction and adduction movements. Reduced individual finger movement is associated with greater hand impairment and the same study also reported unwanted extra finger movements during finger individuation that correlated with lower ARAT and Moberg Pick-Up Test scores.^51^ Hence, finger-tapping that assesses individual movement of the digits is incorporated.

WMFT assesses the functional ability of the UL. Out of the 15 items, 6 components (40%) focus exclusively on the proximal joints. Contrary to that, 3 components of BUFET (25%) assess proximal control, thus ensuring a larger proportion of the scale to focus on diverse hand functions. All the test components of BUFET were administered at ease at the bedside. The BUFET required an average of 10 minutes to complete its administration when compared to the 30–35-minute requirement for WMFT.^13,52^

Our results for the correlation analysis between BUFET and WMFT revealed a high correlation coefficient (r = 0.937, *p*<0.001) which suggest that both tools measure similar, unidimensional construct. The BUFET also demonstrated a high internal consistency with Cronbach’s alpha value of 0.948 (*p*<0.001) which is consistent with the alpha scores of WMFT-0.92,^31^ FMA-U -0.98,^53^ and ARAT -0.98.^54^ The results imply that the BUFET is capable of detecting motor functions nearly identical to the WMFT. In addition to the above, it also suggests that BUFET can be used as an easy-to-administer bedside outcome measure for UL function post-stroke.

### Limitations

Firstly, the items on the scale were narrowed down based on subject experts’ opinions and clinical acumen. An alternative could have been based on direct administration of the components on a limited number of study participants. Secondly, the minimum requirement to carry out the test is that the subject should be made to sit either at the bedside or on a chair which might make its administration difficult for those with a greater degree of motor involvement. Despite employing mini-Delphi method for content validity, our panel’s lack of geographic diversity (South Karnataka-based) may introduce bias. However, the implementation of the mini-Delphi method in the current study was rigorous, adhering to fundamental principles including expert selection, anonymity, iterative consensus rounds, structured communication, and feedback integration.^28,29,55^

### Future scope

Studies should aim at analyzing other psychometric properties including intra-rater and inter-rater reliability based on observations made by multiple observers. The prognostic value of the tool can also be assessed through well-designed prospective studies. A Rasch analysis may help in identifying the key components of the scale to further narrow down the components if indicated.

## Data Availability

The research data associated with this paper and the appendix contents are available from https://doi.org/10.17605/OSF.IO/UFHK5

## Author contributions

DP was the principal investigator (PI) for this study. The conceptualization and the initial development of tool were by the corresponding author (AMJ) who holds the copyright (Reg # L-137026/2023) for the proposed tool. The BUFET is made freely available for teaching and clinical purpose. For research and publication purpose, it is mandatory to credit the corresponding author with relevant citation. Supervision of the trial and the data collection were performed by AN, AJP, and VP. The study was designed, and the data analysis was executed by PM and SA. All the authors equally contributed to the preparation and editing of this manuscript.

## Ethics statement

Approval was obtained from the Research and Institutional Ethics Committee of Kasturba Medical College, Mangalore (IEC KMC MLR 1/2022/15) on 19/01/2022. The study adhered to the ethical principles of the Declaration of Helsinki for research involving human participants.

## Informed consent

Appropriate written informed consent, approved by the Institutional Ethics Committee, was obtained from the study participants. Consent was also taken from the model for animated photographs used in the BUFET description.

## Supporting information

Underlying Data: Bedside Upper Limb Functional Evaluation Tool (BUFET)

## Data availability

### Underlying data

Development and validation of a bedside scale for assessing upper limb function following stroke: A methodological study. DOI: https://doi.org/10.17605/OSF.IO/UFHK5.^56^

This project contains the following data:

- BUFET-Copyright certificate
- BUFET Descriptions
- Data sheet-Concurrent validity; WMFT scores and BUFET scores

Data are available under the terms of the Creative Commons Attribution 4.0 International license (CC-BY 4.0).

## Acknowledgement

Thanks to the Manipal Academy of Higher Education, Manipal for permitting us to carry out this research. Sincere thanks go to all the study participants and special thanks to all the subject experts - Dr. Z.K. Misri, Dr. Shivananda Pai and Dr. Rohit Pai, Department of Neurology, Kasturba Medical College, Mangalore, Dr. Shovan Saha, Department of Occupational Therapy, Manipal College of Health Professions, Manipal and Dr. K Vijaya Kumar and Dr. Shyam Krishnan, Department of Physiotherapy, Kasturba Medical College, Mangalore.

The benefits of publishing with F1000Research:

- Your article is published within days, with no editorial bias
- You can publish traditional articles, null/negative results, case reports, data notes and more
- The peer review process is transparent and collaborative
- Your article is indexed in PubMed after passing peer review
- Dedicated customer support at every stage

For pre-submission enquiries, contact

